# The collagen metabolism, disrupted endothelium, the endothelial progenitor cells and their microvesicles in acute rheumatic fever

**DOI:** 10.1101/2023.08.16.23294151

**Authors:** Sivasubramanian Ramakrishnan, Parul Sahu, Kshama Jain, Jashdeep Bhattacharya, Barun Das, Srikanth Iyer, Anurag Shukla, Arvind Balaji, Saurabh Kumar Gupta, Shyam S Kothari, Anita Saxena, Pramod Upadhay

## Abstract

**Background:** Acute rheumatic fever (ARF) and its chronic sequelae, rheumatic heart disease (RHD) contributes to valvular dysfunction and significant cardiovascular disability and endocardial damage is considered the primary pathophysiological mechanism underlying ARF. This study examined peripheral blood markers of endothelial injury and function in ARF and RHD patients and compared them to healthy controls.

**Method:** In this prospective observational study, the levels of collagen intermediates, matrix metalloproteinases, tissue inhibitors of matrix metalloproteinases, brain natriuretic peptide, Anti-DNaseB, VEGF, E-selectin, VCAM, and ICAM in circulation were estimated. The study also isolated hemangioblastic and monocytic endothelial progenitor cells and their respective microvesicles from the peripheral blood of patients and control samples.

**Results:** Procollagen type I carboxy-terminal propeptide, cross-linked c-telopeptide of type I, and procollagen III c-terminal propeptide levels were higher in RHD subjects compared to patients with ARF. The ARF patients had the highest levels of matrix metalloproteinases 10 (MMP-10) followed by chronic patients and healthy controls. The ratio of tissue inhibitors of matrix metalloproteinases TIMP-1 and MMP-10 was lowest in healthy controls.

At the cellular level, there were higher number of monocytic endothelial progenitor cells (EPCs) in ARF subjects as compared to healthy controls. For hemangioblastic EPCs, there was no significant difference between chronic subjects and healthy controls, though their early subtype was higher in chronic subjects. The hemangioblastic EPCs microvesicles were more abundant in ARF compared to RHD patients.

**Conclusion:** The greater number of EPCs and respective microvesicles confirm the continued disruption of the endothelium in ARF, and during the progression of the disease, the majority of EPCs undergo apoptosis.

**Obituary Statement:** This study was conceived and designed by SR, PU, and Prof. Rajnish Juneja, Professor at AIIMS. Prof. Rajnish Juneja expired in April 2018 while the study was ongoing (1).

Mr. Suran Nambisan, a research fellow at NII, was part of the team who initiated the experimental work. Dr. Suran Nambisan expired in January 2023.

This paper is dedicated to both of them. It was the profound love for mankind and unwavering dedication to perfection by Prof. Rajnish Juneja that brought together a remarkable team to undertake this study. Dr. Suran Nambisan embarked on his professional research journey by successfully establishing and standardizing a few intricate protocols used in this study.

## Introduction

Acute rheumatic fever (ARF) and Rheumatic heart disease (RHD) contribute significantly to cardiovascular morbidity and mortality in low and middle income countries. The disease predominantly affects children and adolescents thereby resulting in substantial loss of disability-adjusted life years. ARF is known to occur as a non-suppurative sequelae to group a beta-hemolytic streptococcal pharyngeal infection.

For a long time, it was believed the molecular mimicry between streptococcal antigens and cardiac proteins can disrupt the immune system, leading to ARF. This mimicry exposes hidden antigens, triggering inflammation and T cell recruitment. This process, called “epitope spreading,” can eventually cause RHD (2-4).

An alternative theory suggests that collagen, not molecular mimicry, may be the target of autoimmunity. It proposes that a complex between streptococcal surface proteins and human type IV collagen could cause an autoantibody reaction and endothelial injury (5).

But there is no conclusive evidence of myocardial necrosis in endomyocardial biopsies of patients, which further suggests that myocardial involvement may not be a contributing factor (6) and the endocardial damage appears to be the primary pathophysiological mechanism by which ARF contributes to valvular dysfunction, leading to various cardiovascular manifestations (7-9).

The objective of the study was to investigate markers of collagen turnover, endothelial activation, and injury. The endothelial injury and repair were investigated by assessing various fractions of endothelial progenitor cells (EPCs). The microvesicles originated from different EPCs were investigated to ascertain their degradation. These parameters were compared among ARF, chronic stable RHD, and age-matched healthy controls.

## Methods

This was a prospective observational study conducted from 2010 to 2020 in the Department of Cardiology, All India Institute of Medical Sciences, New Delhi, and the National Institute of Immunology, New Delhi. Patients aged less than 21 years who were diagnosed with ARF based on the updated Jones criteria were included. Patients with chronic RHD, who were either on medical therapy or awaiting elective cardiac surgery were also enrolled. Diagnosis of RHD was made in all the patients by clinical examination and confirmed by transthoracic echocardiography. The details of confirmation of streptococcus infection and cardiac damage are given in Supplementary Details Section I. Age-matched, with all normal parameters, blood samples were collected from the diagnostic laboratory of AIIMS and were enrolled as healthy controls.

### Ethics Statement

The protocol was approved by the Human Ethics Committee of All India Institute of Medical Sciences, New Delhi (IEC/NP-88/2010, IEC/NP-88/2010, OP-03/09.10.2015, IEC-226/05.04.2019, RP-34/2019) and Institutional Human Ethics Committee of National Institute of Immunology (NII), New Delhi (IHEC#115/19). Written informed consent/assent were obtained from all subjects/guardians before enrolment into the study.

All the experimental work was carried out at NII. All experiments were performed in accordance with National Ethical Guidelines for Biomedical and Health Research Involving Human Participants issued by the Indian Council of Medical Research, Government of India.

### Sample collection

Peripheral venous blood (15 ml) was drawn from all the subjects and the controls, 10 ml in EDTA sprayed K2 vacutainer (BD Biosciences) and was used for peripheral blood mononuclear cells (PBMC) analysis and plasma separation. The other 5ml blood collected in plain vacutainer was used for serum separation. The plasma and serum were stored at -80°C for investigating the selected markers.

Blood samples from 41 patients with ARF and 20 patients with chronic RHD who did not show any evidence of rheumatic activity were collected. Fifteen age matched healthy controls were also enrolled into the study.

A summary of methodology for estimating each of the biomarkers is given below and detailed procedure is given in supplementary details.

Serum titers of collagen intermediates, matrix metalloproteinase–10, tissue inhibitors matrix metalloproteinases, vascular endothelial growth factor (VEGF), brain natriuretic peptide, Anti-DNaseB were measured using their respective commercial kits. Serum concentrations of E-selectin, soluble vascular adhesion molecule 1 (VCAM) and soluble intercellular adhesion molecule 1 (ICAM) were estimated using multiplex bead array kit.

### Endothelial progenitor’s cells isolation and FACS analysis

Peripheral blood mononuclear cells (PBMCs) were isolated from blood and stained with anti-CD14 pecy7 (BD Biosciences, 557742), anti-CD133 APC (BD Biosciences, 566596), anti-309 PE (BD Biosciences, 560494), anti-CD34 FITC (BD Biosciences, 555821). Unstained and single-color controls were prepared for initial voltage settings and fluorochrome compensation. Flowcytometry data acquisition was done using flow cytometer (BD FACSVerse, BD Biosciences) and the data were analyzed using FlowJo v10 software (BD Biosciences).

The hemangioblastic EPCs originate from hemangioblast and these are devoid of any monocytic markers. CD14 marker which is expressed on monocytic EPCs was mainly utilized to distinguish between these two types of EPCs (10).

Hemagioblastic EPCs were further characterized into early and late based on the differential expression of CD133 and CD 309 markers. In order to characterize EPCs, CD34 (a characteristic marker of hematopoietic stem cell), CD133 (expressed on EPCs) and CD309 (expressed in endothelial cells) were used. It has been reported that the expression of CD133 decreases and CD309 increases in the late EPCs (11). The categorization of all the categories of EPCs were performed in all three groups.

### Microvesicles isolation

Microvesicles were isolated from plasma by spinning at 10^5^ g at 4°C for 60 min in an ultracentrifuge. Pellet was washed and resuspended in filtered PBS and stored in -80°C.

### Transmission electron microscopy (TEM)

Microvesicles were subjected to morphological analysis by resuspending small volume of microvesicles on a carbon-coated 200 mesh copper grid photomicrographed under TEM.

### Dynamic light spectroscopy (DLS)

Microvesicles’ size was examined using zetasizer nano zs (Malvern P analytical).

### Staining and analysis of microvesicles

Microvesicles were stained with anti-CD144 APC (Miltenyi Biotec, 130-100-708) and anti-CD31 PE (BD Biosciences, 560983) for 40 min at 4°C. The stained particles were restained with annexin conjugated with FITC (BD Biosciences, 556547). Microvesicles were also stained for endothelial progenitor cells (EPC)-microvesicles and monocytic and monocytic progenitors microvesicles with anti-CD14 V421 (BD Biosciences, 565283), anti-CD133 APC (BD Biosciences, 566596), anti-309 PE (BD Biosciences, 560494), anti-CD34 pecy7 (BD Biosciences, 560710) and annexin conjugated with FITC (BD Biosciences, 556547).

### Microvesicles data acquisition

Microvesicles have size range of 100nm to 1000 nm, which is much smaller than normal cell size. Hence, voltage for scatter plot were set up with the FluoSpheres beads of size 100nm to 1000nm (Flow Cytometry Sub-micron Particle Size Reference Kit, Thermo Fisher Scientific, and F13839). Microvesicles were acquired at the pre-set voltage to ensure that only size-specific MPs are obtained and fluorochrome compensation was done prior to the acquisition of test samples. Data was acquired by flow cytometer (BD FACSVerse, BD Biosciences) and recorded events were analyzed at FlowJo v10 software.

### Total RNA Isolation, cDNA synthesis and qPCR

Total RNA was isolated from blood using Trizol method (Takara, 9108). The genes of MMP-10, MMP-26, MMP-14, MMP-15, MMP-17, MMP-18, MMP-19, MMP-2, MMP-3, MMP-9, MMP-11, MMP-16, MMP-28, MMP-25 were amplified in quantitative real-time PCR with GeneSure™ SYBR Green qPCR Master Mix (2X) (Genetix, PGK025-A) using their respective primers on the Master cycler Real Plex4 platform (Eppendorf, Germany). GAPDH was used as internal control.

### Statistical analysis

Data were expressed in median with interquartile range (IQR) using GraphPad Prism 7 software. Statistical analyses were carried out using non-parametric test; Kruskal–Wallis test and Mann–Whitney U test. For most comparisons p value placed with the respective groups.

## Results

### Baseline parameters

Baseline characteristics of the subjects and the disease-related characteristics are shown in Table 1 and Table 2 respectively. In the ARF group 25 (61%) patients were in NYHA class III or IV and 13 (31.7%) had congestive heart failure at presentation. Among Jones’ major criteria, carditis was seen in 40 (97.6%) patients and joint manifestations in 27 (65.9%) patients. The ASLO levels in ARF and chronic RHD group was 440.1 (±370.9) vs 140.35 (±35.6) respectively (p< 0.001). The number of patients with elevated ESR and CRP in the ARF group was 13 (31.7%) and 23 (56.1%) respectively compared to 5(25%) and 2 (10%) in the chronic RHD group (p 0.001). None of the patients in the chronic RHD group had raised anti-DNAse titres whereas 9 (22%) patients in the ARF group had elevated levels. With regards to underlying cardiac lesion in the ARF group, mixed involvement of mitral and aortic valve was found in 23(6.1%) patients and isolated mitral valve involvement in 17 (41.5%) patients. Isolated mitral involvement was seen in 17 patients (85%) in the chronic RHD group. Isolated aortic valve involvement was seen in only one patient in the ARF group.

**Table 1:** Baseline characteristics of patients.

|  | ARF group<br>(n=41) | Chronic RHD<br>group (n= 20) | p value |
| --- | --- | --- | --- |
| Age (years) * | 12.0 ($\pm 3.1$) | 13.0 ($\pm 3.3$) | |
| Males [n (%)] | 29 (70.7) | 12 (60.0) |  |
| Socio-economic status [n (%)] |  |  |  |
| Lower class | 21 (51.2) | 12 (60.0) |  |
| Lower middle | 17 (41.5) | 6 (30.0) |  |
| Upper middle | 3 (7.3) | 2 (10.0) |  |
| Weight (kg) * | 28.1 ($\pm 7.6$) | 33.5 ($\pm 13.7$) | |
| Height (cm) * | 133.9 ($\pm 12.8$) | 143.70 ($\pm 14.0$) | |
| Body surface area (m<sup>2</sup>) * | 1.03 ($\pm 0.18$) | 1.16 ($\pm 0.25$) | |
| Body Mass Index (kg/m<sup>2</sup>) * | 15.4 ($\pm 2.8$) | 15.9 ($\pm 5.4$) | |
| Systolic blood pressure (mm Hg) * | 107.6 ($\pm 13.6$) | 111.0 ($\pm 14.7$) | |
| Diastolic blood pressure (mm Hg) * | 65.3 ($\pm 12.0$) | 67.8 ($\pm 12.5$) | |
| Pulse rate (beats/min) * | 104.1 ($\pm 17.4$) | 91.5 ($\pm 17.2$) | |
| NYHA class [n (%)] |  |  |  |
| IV | 9 (22.0) | 0 |  |
| III | 16 (39.0) | 0 |  |
| II | 14 (34.1) | 8 (40) |  |
| I | 2 (4.9) | 12 (60) |  |
| Congestive heart failure [n (%)] | 13 (31.7) | 0 |  |
| Six-minute walk distance ** | 350.0 (326.8 – 375.0) | 425.0 (404.0 – 448.0) | 0.807 |
| Previous history of ARF [n (%)] | 14 (34.1) | 3 (15) |  |
| Getting secondary prophylaxis [n (%)] | 18 (43.9) | 14 (70) |  |
| Hemoglobin (g/dL) * | 10.7 ($\pm 1.5$) | 11.6 ($\pm 1.7$) | |

\*Values expressed as mean (standard deviation)
\*\* Values expressed as Median (interquartile range)
NYHA: New York Heart Association.

**Table 2:**
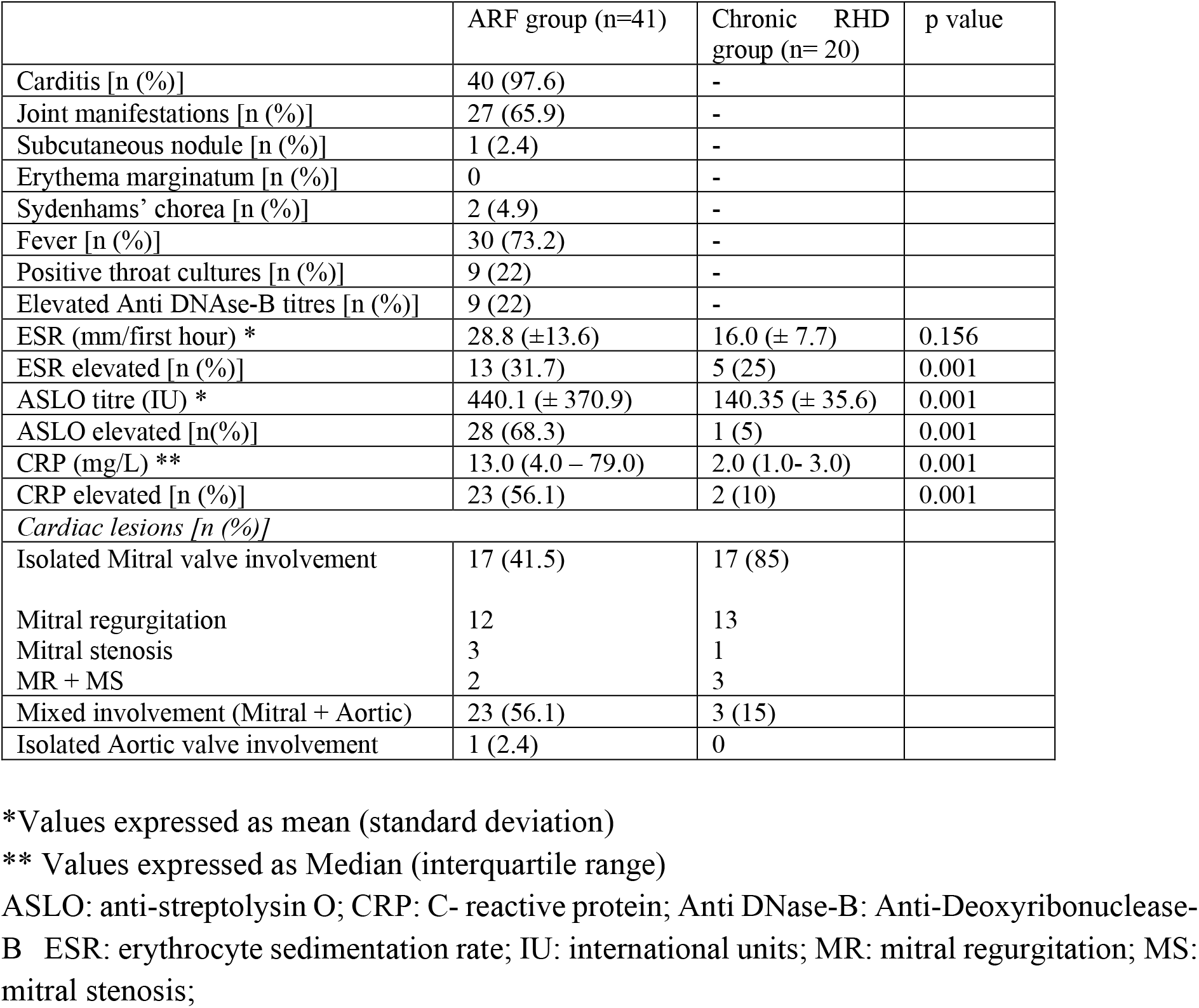
Disease related characteristics (clinical and laboratory) of patients.

### Parameters of collagen turnover

#### Components of extracellular matrix remodeling in ARF and Chronic RHD

The components involved in the metabolism of type I and type III collagen were estimated. Procollagen type I carboxy-terminal propeptide (PICP) released during the synthesis of type I collagen was found to be higher in chronic RHD subjects compared to patients with ARF. Both these groups had significantly higher PICP levels as compared to controls. Cross-linked c-telopeptide of type I collagen (CTXI) which is released in the blood during degradation of type1 collagen was higher in the ARF group compared to chronic with (*p* 0.001). The titres were significantly higher in both the chronic RHD group and ARF group compared to healthy controls. The ratio of PICP/CTXI was calculated to assess collagen turnover.

In ARF patients, the ratio was [Median (IQR ng/ml)] 0.7392 (0.6809-0.7817) versus 0.9259 (0.8617-0.9608) in chronic RHD patients (P 0.0001). Titers of procollagen III c-terminal propeptide (PIIICP) were higher in ARF subjects as compared to the chronic subjects (p <0.0001) (Table 3).

**Table 3:** Titres of intermediates of collagen turnover in each group of subjects.

| Markers | Healthy control<br>(n=10) | ARF group<br>(n=41) | Chronic RHD group<br>(n=20) | P value<br>(ARF vs<br>RHD) | P value<br>(ARF vs<br>Healthy) | P value (RHD<br>vs Healthy) |
| --- | --- | --- | --- | --- | --- | --- |
| PICP<br>(pg/ml) | 65456<br>(55732 - 78274) | 70322<br>(60359 - 78481) | 80488<br>(73466 - 82732) | 0.0017 | 0.4326 | 0.0303 |
| CTXI<br>(ng/ml) | 0.1173<br>(0.1089 - 0.1377) | 2.313<br>(2.25 - 2.41) | 2.244<br>(2.169 - 2.483) | 0.0013 | < 0.0001 | <0.0001 |
| PIIICP<br>(pg/ml) | 98.49<br>(82.8 - 153.5) | 89.97<br>(87.9 - 97.8) | 85.71<br>(82.9 - 88.38) | <0.0001 | 0.7747 | 0.1863 |
| PICP/CTXI | 0.6706<br>(0.5 - 0.7945) | 0.7392<br>(0.6809 - 0.7817) | 0.9259<br>(0.8617 - 0.9608) | < 0.0001 | 0.193 | 0.0003 |

Matrix metalloproteinases are involved in the degradation of proteins of the extracellular matrix. MMPs are classified into 24 categories. To identify the best MMP as a diagnostic marker, the whole blood mRNA expressions of different MMPs were quantified and compared among the selected subgroups (healthy=3, ARF=3, Chronic=3) (Supplementary details, section III). The MMP-10 expression was significantly higher in chronic subjects with *p*=0.0017 compared to healthy controls. The titre of MMP-10 in ARF patients was [Median (IQR)] 152762 (109436 - 178048) pg/ml compared to 112351 (91434 - 132402) pg/ml in chronic RHD subjects (p 0.0005).

The functional activity of matrix metalloproteinases is regulated by tissue inhibitors of matrix metalloproteinases (TIMPs) and they play a critical role in maintaining the balance of tissue degradation. The titres of all four classes of TIMP were significantly higher in the ARF patients compared to chronic RHD patients (p <0.001) as shown in Table 4.

**Table 4:** Titres of tissue inhibitors of matrix metalloproteinases in each group of subjects.

| Markers | Healthy control<br>(n=10) | ARF group<br>(n=41) | RHD group<br>(n=20) | P value (ARF vs<br>RHD) | P value (ARF<br>vs Healthy) | P value<br>(RHD vs<br>Healthy) |
| --- | --- | --- | --- | --- | --- | --- |
| TIMP-1<br>(ng/ml) | 101<br>(90.3 - 112.2) | 143.6<br>(133.9 - 153.6) | 113.6<br>(107.3 - 130.3) | 0.0001 | 0.0001 | 0.0108 |
| TIMP-2<br>(ng/ml) | 2.145<br>(0 - 18.3) | 47.06<br>(9.89 - 140) | 9.858<br>(0 - 47.05) | 0.001 | 0.0004 | 0.1168 |
| TIMP-3<br>(pg/ml) | 1812<br>(1634 - 3673) | 6656<br>(2713 - 33234) | 2218<br>(1832 - 2635) | 0.0002 | 0.0004 | 0.2865 |
| TIMP-4<br>(pg/ml) | 3517<br>(1864 - 9684) | 17425<br>(8116 - 61401) | 6491<br>(3724 - 8367) | 0.0003 | 0.0005 | 0.3021 |

Since TIMP-1 and TIMP-2 are known to interact with MMP-10 (12), the ratio of MMP-10/TIMP-1 and MMP-10/TIMP-2 were calculated. The ratios were compared within the ARF, chronic and healthy subjects, these were significantly higher in chronic subjects as compared to ARF (*p* =0.0051) and healthy (*p*=0.0014) group. (Table 5)

**Table 5:**
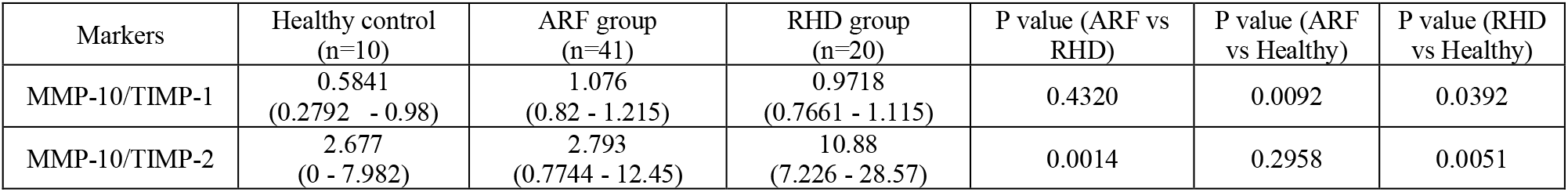
Ratio of MMPs and TIMPs.

### Endothelial injury and repair

Hemangioblastic EPCs are actual precursors of endothelial cells, while monocytic EPCs are the ‘helper cells’ that aid hemangioblastic EPCs to get recruited at the site of damage by releasing angiogenic factors (10, 13). Therefore, a comparison of hemangioblastic EPCs (CD14^-^ CD133^+^ CD34^+^ CD309^+^) and monocytic EPCs (CD14^+^ CD133^+^ CD34^+^ CD309^+^) was done. In ARF and chronic subjects, higher level of monocytic EPCs compared to Hemangioblastic EPCs were present with *p* = 0.0384 and *p* = 0.0521 respectively (Figure 1). Among the healthy controls this difference was not significant with p = 0.093.

**Figure 1.**
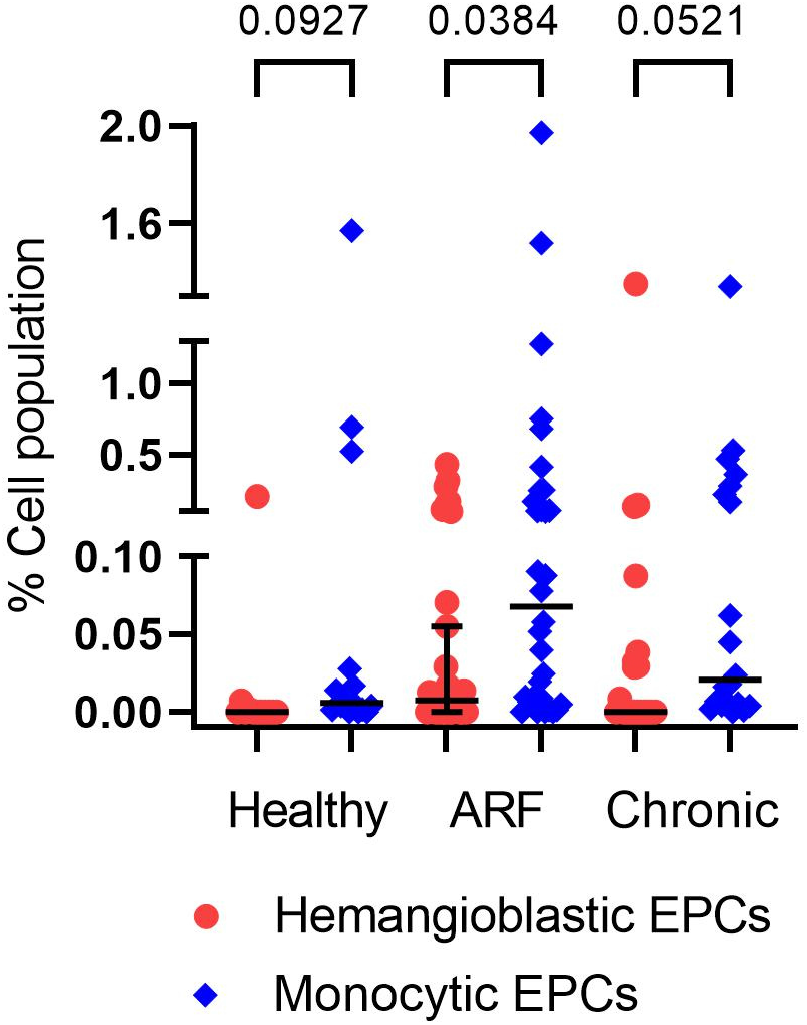
The comparison of different EPCs.

#### EPCs subsets

To investigate the damage of endothelial lining, EPCs from the PBMCs of ARF, chronic RHD and healthy subjects were investigated by flowcytometery. The gating strategy for all the EPCs subsets and representative graphs is shown in Supplementary details section IV and results are summarized in Figure 2.

**Figure 2.**
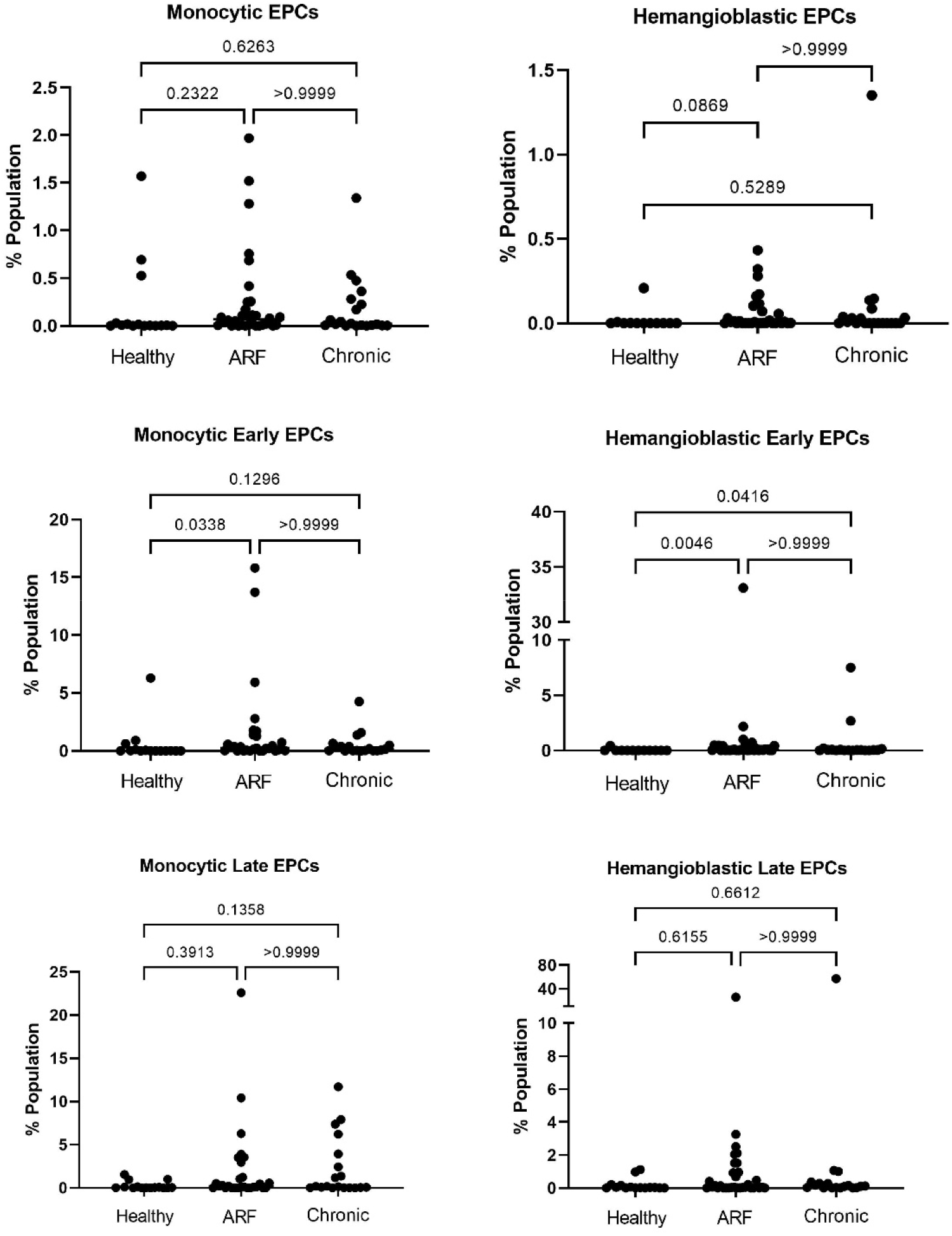
Monocytic and Hemangioblastic EPCs in different groups.

In case of early monocytic EPCs, higher percentages of EPCs were observed in ARF subjects as compared to healthy controls with *p*=0.0338. Similarly, higher EPCs were present in chronic subjects with *p*=0.129 compared to healthy controls. Likewise, late monocytic EPCs were identified, in which the higher percentages of EPCs were observed in chronic subjects compared to healthy controls with *p*=0.135 (Figure 2).

There were significantly higher hemangioblastic EPCs in the ARF compared to healthy (p=0.086), and like the monocytic EPCs, hemangioblastic EPCs percentages were similar among other comparisons. The early hemangioblastic EPCs were higher both in ARF as well as chronic subjects compared to healthy controls with p=0.0046 and p= 0.041 respectively. ARF and chronic both has similar percentages of early hemangioblastic EPCs. The late hemangioblastic EPCs percentages were similar among all comparisons (Figure 2).

#### Endothelial specific microvesicles

Microvesicles are the class of vesicles released by cells that are under stress or undergoing apoptosis or necrosis. These vesicles are identified by markers of the parental cells. Confirmation of size and morphology of microvesicles were done by transmission electron microscopy (TEM) and dynamic light scattering (DLS) and details are given in Supplementary details Section V, Figure S4a and S4b.

To evaluate the damage of endothelium, endothelial cells specific microvesicles were investigated. There were more Hemangioblastic EPCs microvesicles and Hemangioblastic early EPCs microvesicles in ARF compared to the healthy controls. Data are summarized in Fig. 3 and Supplementary details, Figure S4c and S4d.

**Figure 3.**
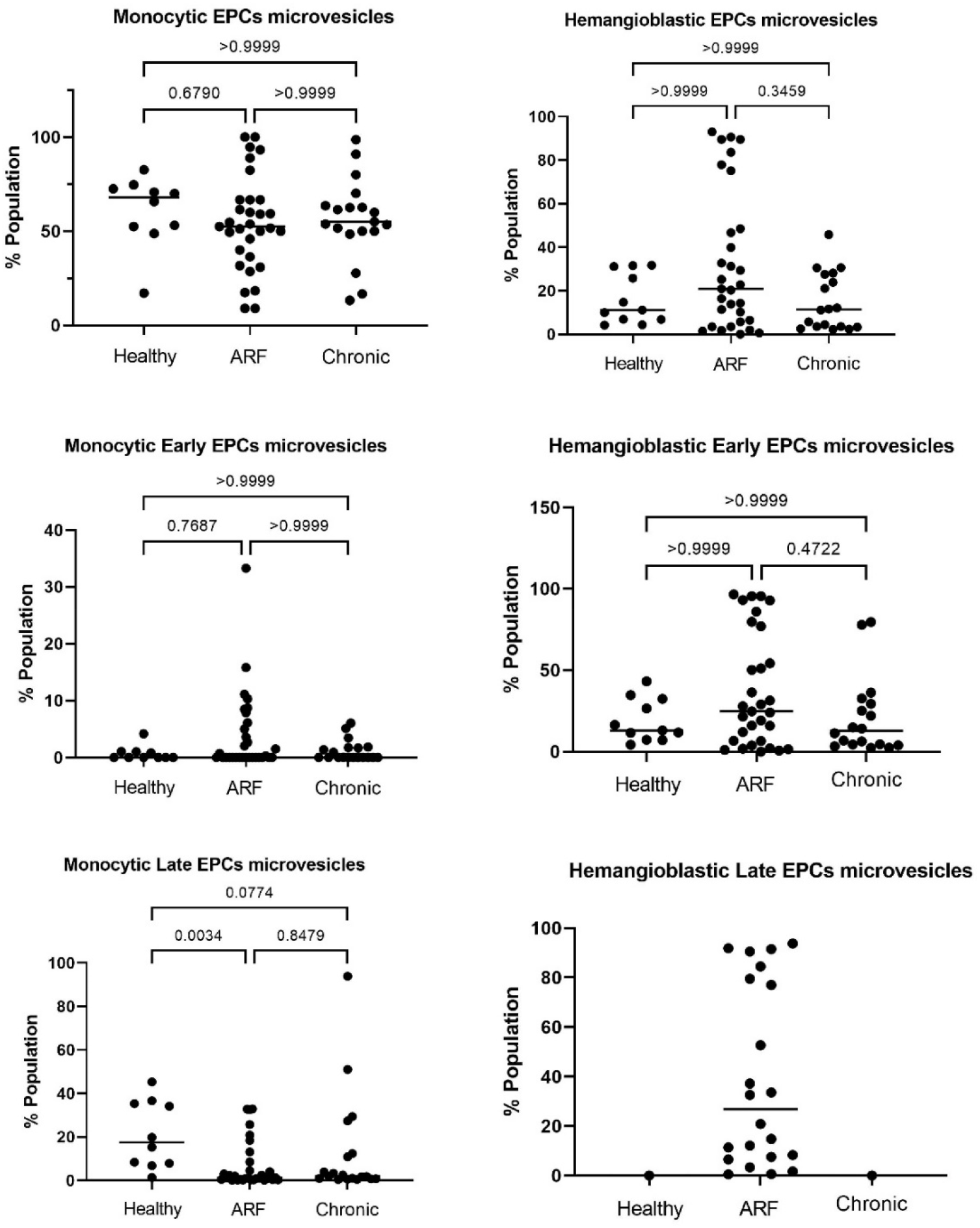
The Microvesicles of Monocytic and Hemangioblastic EPCs in different groups.

#### Neo-angiogenesis and inflammation

VEGF is a known factor for Neo-angiogenesis and EPCs mobilization (14). VEGF levels were not significantly different between healthy and ARF subjects but it was lower in chronic RHD compared to ARF subjects (*p* 0.0311) (Figure 4A).

**Figure 4.**
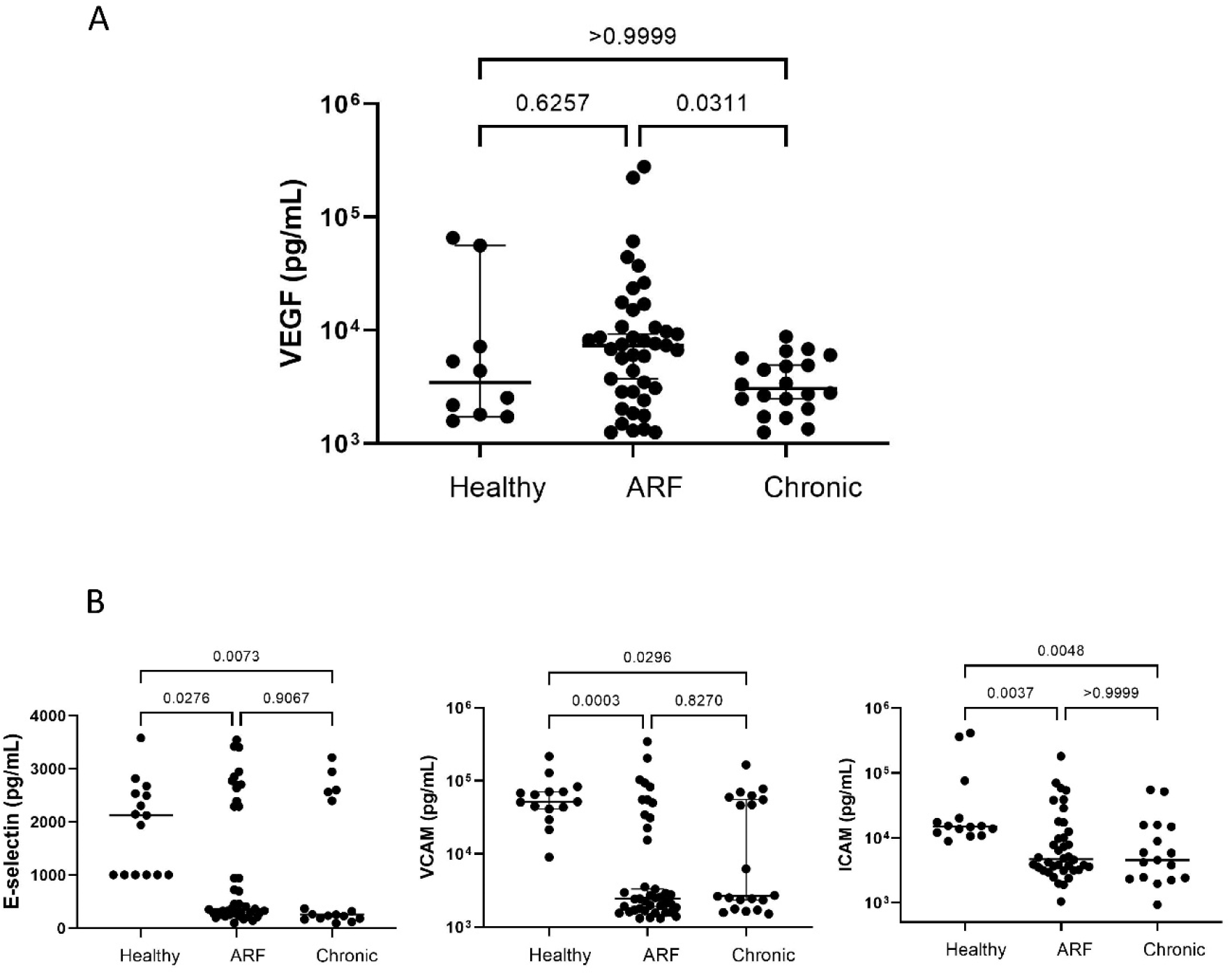
Neo-angiogenesis and inflammation. (A) VEGF levels and (B) adhesion molecules in different groups.

Adhesion molecules such as vascular cell adhesion molecule (VCAM), intercellular adhesion molecule (ICAM) and E-selectin are involved in the transendothelial migration of leukocytes (15). Contrary to the trends typically reported (15-17), lower levels of E-selectin, VCAM, and ICAM were found in ARF and chronic compared to healthy subjects (Figure 4B).

## Discussion

Since the initial understanding of acute rheumatic fever has been commonly defined by the occurrence of hyperglobulinemia, lymphadenopathy and splenomegaly, along with lesions affecting mesenchymal tissue, the heart, blood vessels, joints, and serous membranes. This pattern of multi-system involvement is typical of collagen diseases (18).

In this study the metabolism of collagen was comprehensively investigated. The findings corroborated well with Tanima et.al. who reported the change in collagen metabolism markers in rheumatic mitral stenosis (MS) and mitral regurgitation (MR). They have shown altered levels of PICP, PIIINP, MMP-1 and TIMP-1 in chronic RHD cases as compared to healthy controls (19).

Similar observations were made by Sarkar et.al. that PICP levels were elevated in ARF and chronic cases (20). C-terminal propeptide of collagen alpha-1(I) chain (PICP) is released during the synthesis of type I collagen (21). The values were found to be significantly higher in chronic RHD subjects as compared to ARF and healthy controls in this investigation.

The raised PICP titers during the progression of ARF to chronic stages suggest that the synthesis of collagen increases in the later stage where the valvular fibrosis and scarring is expected to occur.

In addition, it was found that CTXI levels were significantly higher in both the ARF and chronic groups, compared to healthy controls. CTXI is a marker of degradation of type 1 collagen. The CTXI levels were significantly higher in the ARF subjects compared with chronic subjects. This suggests that CTXI could be a potential biomarker candidate to distinguish between active and chronic stages of the disease.

This also reaffirms that the type 1 collagen degradation may be happening early during occurrence of carditis (22, 23). Whereas, collagen turnover was noticed to be higher in the chronic group compared to ARF group, as shown by a higher ratio of PICP to CTXI in the chronic group again supporting the hypothesis that valve remodeling continues to occur even in during the stable phase of the disease process. Further, an intermediate of type III pro-collagen (PIIICP) was investigated and significantly higher levels were observed in ARF as compared to chronic subjects which confirm that excess of this collagen is synthesized during early stage of chronic.

Next, the potential inflammation arising from the degradation of collagen was investigated. During acute inflammation there is an activation of fibroblast and endothelial cells (24). These activated cells release MMPs, which clears off the deposited collagen (25), thus different classes of MMPs and TIMPs which inhibit MMPs, were estimated in ARF, chronic subjects and healthy controls. Further the MMP/TIMP ratio was calculated to understand their interactions (12).

This ratio was higher in chronic subjects compared to ARF cases. The higher ratio indicates that MMP-10 synthesis was elevated in chronic subjects relative to TIMP-2. This confirms that a greater amount of collagen was degraded among chronic cases. Additionally, it suggests that MMP-10 levels can be used to differentiate between ARF and the chronic stage of the disease.

MMPs also play an important role in Neo-angiogenesis. MMPs break down fibrillar collagen or proteoglycan proteins, facilitating the migration of endothelial cells and the formation of new vessels (26, 27).

Neo-angiogenesis further promote inflammation by enabling immune cells and soluble inflammatory factors to enter the valvular. Secondly, the development of vascular networks can contribute to the deterioration of valve tissue by disrupting its natural architecture (28, 29).

The neo-angiogenesis was accessed by measuring the levels of vascular endothelial growth factor (VEGF). There was no significant difference in VEGF levels in ARF and chronic groups compared to the healthy controls.

Cell adhesion molecules are important components of an active T-cell mediated immune response (10). Lower levels of adhesion molecules; E-selectin, VCAM and ICAM were observed in ARF and chronic patients compared to healthy controls.

These lower levels contradict the findings of the inflammation in streptococcal sequelae due to the autoimmune mechanism resulting from the molecular mimicry between streptococcal M protein antigens, cardiac myosin, the cross-reactivity due glycosylated proteins such as laminin or a few extracellular proteins (30, 31).

Additionally, this observed trend is opposite of a few earlier reports in which inflammation in the valve endothelium resulted in the higher expression of vascular cell adhesion molecules (32) and elevated levels of E-selectin, VCAM and ICAM were reported in ARF compared to healthy controls (16, 17).

Although anomalous, the lower levels of cell adhesion molecules corroborate well with the similar VEGF levels in all the groups. Secondly, the effect of previous or ongoing antibiotics treatment perhaps has some role in normalizing these levels.

Further, the response arising from the fragmented endothelium was examined. The cascade of events following injury to endothelial cells leads to the release of Endothelial Progenitor Cells (EPCs) from the bone marrow into circulation (10).

Similar trend was observed in ARF and chronic sets, and the difference between numbers of monocytic EPCs and hemangioblastic EPCs became more pronounced in ARF and chronic sets (Figure 1) indicating deeper involvement of EPCs in the progression of the disease.

The scenario becomes clearer when various subsets of monocytic and hemangioblastic EPCs were examined. Significantly higher numbers of hemangioblastic early EPCs and monocytic early EPCs reaffirms the initiation of repair mechanism of disrupted endothelium. Although, the differences were less significant in late EPCs subsets.

It could be hypothesized that in order to facilitate the homing of hemangioblastic early EPCs to the site of repair, the circulatory system elevates the levels of monocytic early EPCs in response to endothelial damage.

In the real-life scenarios of the disease, despite the presence of circulatory EPCs, the transition from ARF to a chronic condition often persists due to the continues damage inflicted on the endothelium. In order to explore the reparative potential of EPCs, the cellular viability of different EPCs subtypes was evaluated through the quantification of their respective microvesicles. Microvesicles are a type of extracellular vesicle that are released by cells during the pre-apoptotic stage (33).

Except in the hemangioblastic EPCs microvesicles and its early subset, the percentages of microvesicles were similar in all the groups. Between the ARF and chronic, though the percentage of hemangioblastic EPCs were similar, there were more hemangioblastic EPCs microvesicles in the ARF compared to the chronic group. This is another indication that in ARF, majorly the EPCs were undergoing apoptosis and the endothelium continues to remain disrupted.

## Supporting information

Supplementary Details

## Data Availability Statement

All relevant data is contained within the article and supplementary details.

## Acknowledgement

We are grateful to all the patients, healthy controls, and their guardians for their participation in the study.

## Authors contributions

SR, Rajnish Juneja and PU conceived and designed the study and analysed data. Initial standardization was done by JB, BD, SI, Suran Nambisan and PU. Majority of the samples were processed by PS and KJ. Isolation and characterization of microvesicles was done by PS and AS. Patients were diagnosed and enrolled by SR, Rajnish Juneja, and AB. Healthy controls were enrolled by SR and AB. The first draft of the manuscript was written by SR, AB, and PS, and editing was done by PU. All authors read and approved the final manuscript.

