## Supplementary Details for "The collagen metabolism, disrupted endothelium, the endothelial progenitor cells and their microvesicles in acute rheumatic fever"

Sivasubramanian Ramakrishnan[1#],
Parul Sahu[2],
Kshama Jain[2],
Jashdeep Bhattacharya[2],
Barun Das[2],
Srikanth Iyer[2],
Anurag Shukla[2],
Arvind Balaji[1],
Saurabh Kumar Gupta[1],
Shyam S Kothari[1],
Anita Saxena[1],
Pramod Upadhay[2#],

[1] Department of Cardiology, All India Institute of Medical Sciences, New Delhi
[2] National Institute of Immunology, Aruna Asaf Ali Marg, New Delhi

#### Obituary Statement

This study was conceived and designed SR, PU and Prof. Rajnish Juneja, Professor at AIIMS, while the study was undergoing, Prof. Rajnish Juneja expired in April 2018. Mr. Suran Nambisan, a research fellow at NII, was part of the team who initiated experimental work. Dr. Suran Nambisan expired in January 2023. This paper is dedicated to both of them. It was the profound love for mankind and unwavering dedication to perfection by Prof. Rajnish Juneja that brought together a remarkable team to undertake this study. Dr. Suran Nambisan embarked on his professional research journey by successfully establishing and standardizing a few intricate protocols used in this study.

#### Index

| | page |
|---|---|
| Detailed Methods | 2 |
| Confirmation of streptococcus infection and cardiac damage in selected subjects | 6 |
| Gene expression analysis of MMPs | 7 |
| Gating strategy for all the EPCs subsets and a representative graph | 8 |
| Size, morphology and percentage of microvesicles | 9 |
| Enumeration of monocyte microvesicles with their specific cell types | 10 |
| Gating strategy for EPC microparticles | 11 |

**Detailed Methods**

This was a prospective observational study conducted from 2010-2020 in Department of Cardiology, All India Institute of Medical Sciences, New Delhi and National Institute of Immunology, New Delhi. We included patients aged less than 21 years, who were diagnosed with ARF based on the updated Jones criteria. We also enrolled patients with chronic RHD, who were either on medical therapy or awaiting elective cardiac surgery. Diagnosis of RHD was made in all the patients by clinical examination and confirmed by transthoracic echocardiography. Age matched healthy controls were also enrolled into the study.

**Ethical Statement**

The protocol was approved by the human ethics committee of All India Institute of Medical Sciences (ref no: IEC/NP-88/2010). After initiation of the study, additional markers were included after appropriate intimation and approval of the committee (ref no IEC/NP-88/2010, OP-03/09.10.2015). The protocol was modified to include healthy controls after approval by the human ethics committees of All India Institute of Medical Sciences, New Delhi (ref no: IEC-226/05.04.2019, RP-34/2019) and National Institute of Immunology, New Delhi (ref no IHEC#115/19). Written informed consent/assent were obtained from all subjects and guardians before enrolment into the study.

**Sample collection**

Peripheral venous blood (15 ml) was drawn from all the subjects and the controls, 10 ml in EDTA sprayed K2 vacutainer (BD Biosciences) and was used for peripheral blood mononuclear cells (PBMC) analysis and plasma separation. The other 5ml blood collected in plain vacutainer was used for serum separation. The plasma and serum were stored at -80◦C for investigating the selected markers. The laboratory methodology for estimating each of the biomarkers is described below.

**Enzyme linked immunosorbent assays (ELISA)**

Serum titers of PICP, CTXI, PIIICP, TIMP-1, TIMP-2, TIMP-3, TIMP-4, VEGF, MMP-10, anti-DNase and BNP were measured using their respective commercial kits.

**Measurement of collagen intermediates**

Serum samples were used for the estimation of titers of various collagen intermediates using commercially available kits (E-Lab sciences) following manufacturer instructions. Serum dilutions were made 1/8 and measurements of procollagen type I carboxy-terminal propeptide (PICP) (E-Lab sciences, E-EL-H0196), cross-linked c-telopeptide of type I collagen (CITP) (E-Lab sciences, E-EL-H0835) and procollagen III c-terminal propeptide (PIIICP) (E-Lab sciences, E-EL-H0841) were performed.

**Measurement of Matrix metalloproteinase - 10 (MMP- 10)**

Stored serum aliquots were thawed and diluted 1/16 as suggested by the manufacturer (E-Lab sciences, E-EL-H0132).Briefly, standards were prepared by reconstituting the lyophilized vial. Standards were prepared in the range of 2500 µg/ml to 0µg/ml. Subsequently, serum was incubated in precoated anti-MMP-10 wells along with standards and blanks for 90 min at 37˚C. Thereafter, liquid was aspirated and wells were again incubated with the biotinylated antibody for 1hr at 37˚C. Wells were then washed with buffer provided along with the kit and were incubated with HRP conjugated antibody for 30 min at 37˚C. Wells were washed again and incubated with a substrate solution followed by a stop solution. The absorbance was recorded at 450nm and standard graph was plotted against absorbance and concentration.

**Estimation of tissue inhibitors matrix metalloproteinases (TIMPs)**
TIMPs were measured in the serum samples with 1/8 dilution. Precoated ELISA plates from E-Lab sciences for TIMP-1 (E-Lab sciences, E-EL-H0184), TIMP-2 (E-Lab sciences, E-EL-H1453), TIMP-3 (E-Lab sciences, E-EL-H1454) and TIMP-4 (E-Lab sciences, E-EL-H1455) were used.

**Measurement of vascular endothelial growth factor (VEGF)**
VEGF levels were estimated using ELISA kit from Invitrogen (BMS277-2). 12.5 µl of the accompanying capture antibody was diluted to 10 ml in the coating buffer. Each well of a 96-well flat bottom high protein binding plate (Maxisorb, Nunc, USA) was coated with 100 µl of diluted capture antibody solution at 4˚C overnight. The plate was then washed thrice with a wash buffer and wells were blocked with 200 µl of 1X assay diluent (eBioscience, USA) for 60 min at 25˚C. Wells were washed again thrice with wash buffer. VEGF standard dilutions from 1500 pg/ml to 0 pg/ml were prepared using assay diluents and 100 µl of standard and sample was added to each well and incubated for 2 hrs at 25˚C. After incubation, wells were washed with washing buffer. 100 µl of a solution prepared by diluting 1.25 µl of the provided detection antibody diluted to 10 ml using the assay diluents was added to each well and incubated at 25˚C for 1 hour. This was followed by three washes and addition of 100 µl of a solution prepared by diluting 2.5 µl of streptavidin HRP to 10 ml using assay diluent was added to each well and incubated at 25 ˚C for 30 minutes. Then wells were washed for 5 times with wash buffer and 100 µl of 3,3′,5,5′-tetramethylbenzidine (TMB) substrate was added to each well and incubated until the color changed to blue or for a maximum for 5 min and further change in colour was stopped by addition of 50 µl of 2N $H_2SO_4$. Addition of stop solution changed the blue color of this solution to yellow after which it was read at 450 nm using an ELISA plate reader. The standard graph was plotted against absorbance and concentration. The results were expressed in pg/ml.

**Measurement of Brain natriuretic peptide (BNP)**
Immunological assay of serum BNP titers were performed on commercially available ELISA kit (Cloud clone, CEA541Hu) following manufacturers's direction.

**Measurement of Anti-Deoxyribonuclease B Antibody (Anti-DNaseB)**
Titers of anti- DNase B were determined using commercially available Anti-DNaseB ELISA kit (Cloud clone, AEA311Hu) . All the procedures were followed as per the manufacturer's instructions.

**Multiplex bead array**
Serum concentrations of E-selectin, soluble vascular adhesion molecule 1 (sVCAM) and soluble intercellular adhesion molecule 1 (sICAM) were identified using multiplex bead array kit (NR basic kit, aimplex, P100001) with analytes E-selectin (aimplex, A113254), sVCAM (aimplex, A113311) and sICAM (aimplex, A113263) and the procedure was performed as per the given protocol. Stained samples were acquired by flow cytometer (BD FACSVerse, BD Biosciences). The acquired data were analyzed using FCAP Array Software v3.0 (BD Biosciences).

**Endothelial progenitor's cells isolation and FACS analysis**
10 ml of blood was diluted with RPMI up to 4.4 X dilution and layered on HiSep™ LSM 1077, then centrifuged at 400g, 25˚C for 30 min. Thereafter, buffy coat containing PBMCs were washed with phosphate buffer saline (PBS). PBMCs were stained with anti-CD14 pecy7 (BD

Biosciences, 557742), anti-CD133 APC (BD Biosciences, 566596), anti-309 PE (BD Biosciences, 560494), anti-CD34 FITC (BD Biosciences, 555821). Unstained and single-color controls were prepared for initial voltage settings and fluorochrome compensation. Concentration of antibodies was used as per given protocol. All the stained samples were incubated at 4˚C for 40 min and subsequent washes with PBS were done to remove any unbound antibodies. Cells were then fixed at 1X lyse/Fix buffer (Lyse/Fix Buffer 5X BD Phosflow™, BD Biosciences, 558049) in the dark for 15 min at 25˚C and then washed with PBS. Sample acquisition was done using flow cytometer (BD FACSVerse, BD Biosciences) and the data were analyzed using FlowJo v10 software (BD Biosciences).

**Microparticle isolation**

Stored plasma was thawed at 25°C and cellular debris were removed by spun in the centrifugation at 2000g, 4°C for 30 min. Then supernatant was centrifuged at 1 lakh g, 4°C for 60 min in the ultracentrifuge. Pellet was washed with filtered phosphate buffer saline (PBS) (1 lakh g, 4°C for 60 min in ultracentrifuge). Finally, pellet was resuspended in filtered PBS and stored in -80°C.

**Transmission electron microscopy (TEM)**

Microvesicles were subjected to morphological analysis by resuspending small volume of microvesicles on a carbon-coated 200 mesh copper grid (PELCO® TEM, Ted Pella, INC.) for 1 min. Thereafter, sample was stained with uranyl acetate 30% for 30 seconds and washed with double distilled water. Copper grid was dried and photomicrographed under TEM.

**Dynamic light spectroscopy (DLS)**

Microvesicles' size was examined using zetasizer nano zs (Malvern P analytical). Isolated microvesicles were resuspended in 1ml of filtered PBS and sizes of different particles were evaluated when the laser strikes on the suspended particles. The scattering patterns were plotted against the size distribution by intensity graph. The graph obtained confirmed the size of different types of particles in the solution.

**FACS staining and analysis of microvesicles**

Isolated microvesicles were resuspended in 1X annexin binding buffer (BD Biosciences, 556454) and redistributed in equal volumes to different micro-centrifuge tubes. For enumerating endothelial microvesicles, these particles were stained with anti-CD144 APC (Miltenyi Biotec, 130-100-708) and anti-CD31 PE (BD Biosciences, 560983) for 40 min at 4°C. The stained particles were restained with annexin conjugated with FITC (BD Biosciences, 556547) and incubated at 25°C for 15 min. Similarly, particles were also stained for endothelial progenitor cells (EPC)-microvesicles and monocytic and monocytic progenitors microvesicles with anti-CD14 V421 (BD Biosciences, 565283), anti-CD133 APC (BD Biosciences, 566596), anti-309 PE (BD Biosciences, 560494), anti-CD34 pecy7 (BD Biosciences, 560710) and annexin conjugated with FITC (BD Biosciences, 556547).

**Microparticle data acquisition**

Microvesicles are vesicles within a size range of 100nm to 1000 nm, which is much smaller than normal cell size. Hence, voltage for scatter plot were set up with the FluoSpheres beads of size 100nm to 1000nm (Flow Cytometry Sub-micron Particle Size Reference Kit, Thermo Fisher Scientific, and F13839). Log SSC and linear FSC plot were used to acquire the beads. P1 gate was assigned to this specific size range and the number of acquisition events was defined. Microvesicles (MVs) were acquired at the pre-set voltage to ensure that only size-specific MPs are obtained. Electronic background was excluded using unstained and single colour control and fluorochrome compensation was done prior to the acquisition of test samples. The next plot was designed as FSC vs. annexin fluorochrome (FITC) and only P1 gate acquired data was recorded. All the annexin$^{+}$ events were gated as P2 and enumerated as microvesicles. The samples were acquired by flow cytometer (BD FACSVerse, BD Biosciences) and recorded events were analyzed at FlowJo v10 software. Similarly, different cell-specific microvesicles were analyzed by the above-mentioned strategy and using P1 (size-specific) and P2 (Annexin$^{+}$) as initial gates.

**Total RNA Isolation, cDNA synthesis and qPCR**

Total RNA was isolated from 250µl of blood using Trizol method (Takara, 9108). Protocol was used as per the direction of manufacturer. Purity and concentration of RNA were checked in spectrophotometer (Nanodrop 2000, Thermo scientific USA). RNA with absorbance (260/280) of more than 1.8 were selected and complementary DNA prepared using iScript cDNA synthesis kit (Bio Rad, 1708891) as per manufacturer instructions. Then, genes of MMP-10, MMP-26, MMP-14, MMP-15, MMP-17, MMP-18, MMP-19, MMP-2, MMP-3, MMP-9, MMP-11, MMP-16, MMP-28, MMP-25 were amplified in quantitative real-time PCR with GeneSure™ SYBR Green qPCR Master Mix (2X) (Genetix, PGK025-A) using their respective primers on the Master cycler Real Plex4 platform (Eppendorf, Germany). GAPDH was used as internal control.

**Statistical analysis**

Comparisons were made among all the three groups. All the data were expressed in median with interquartile range (IQR) using GraphPad Prism 7 software. Statistical analyses were carried out using non-parametric test; Kruskal–Wallis test and Mann–Whitney U test. For multiple comparisons Dunn's Test were used. $P < 0.05$ was considered as statistically significant.

**Confirmation of streptococcus infection and cardiac damage in selected subjects**

The infection of streptococcus was confirmed by the titre of anti-DNase B antibodies. A significant increase of titres in diseased subjects as compared to healthy controls indicates an infection of streptococcus in ARF and chronic subjects (Figure S1a).

To determine the cardiac damage, brain natriuretic peptide (BNP) was utilized. The normal threshold for the BNP level is 35pg/ml. The BNP level was estimated in ARF, chronic and healthy subjects. The majority of ARF subjects were above the 35pg/ml with $p$=0.0227 as compared to healthy controls. However, 99% of selected chronic subjects titres were more than 35pg/ml and were higher with $p$=0.0011 as compared to healthy subjects. This indicates that the selected diseased subjects have cardiac damage.

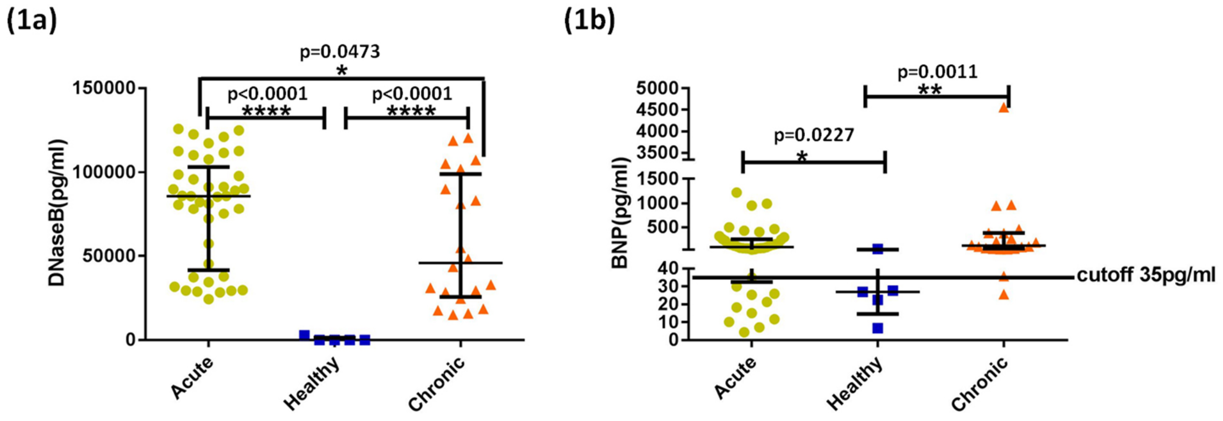


*Figure S1. Estimation of Streptococcus infection and cardiac damage in the ARF, chronic and healthy subjects. (a) Anti-Deoxyribonuclease B (DNaseB) titer and (b) Brain natriuretic peptide (BNP) titer. The 35pg/ml is the cutoff for BNP. Data are median with bars representing an interquartile range (IQR).*; p<0.05. ARF=41, chronic=20 and healthy=5*

### Gene expression analysis of MMPs

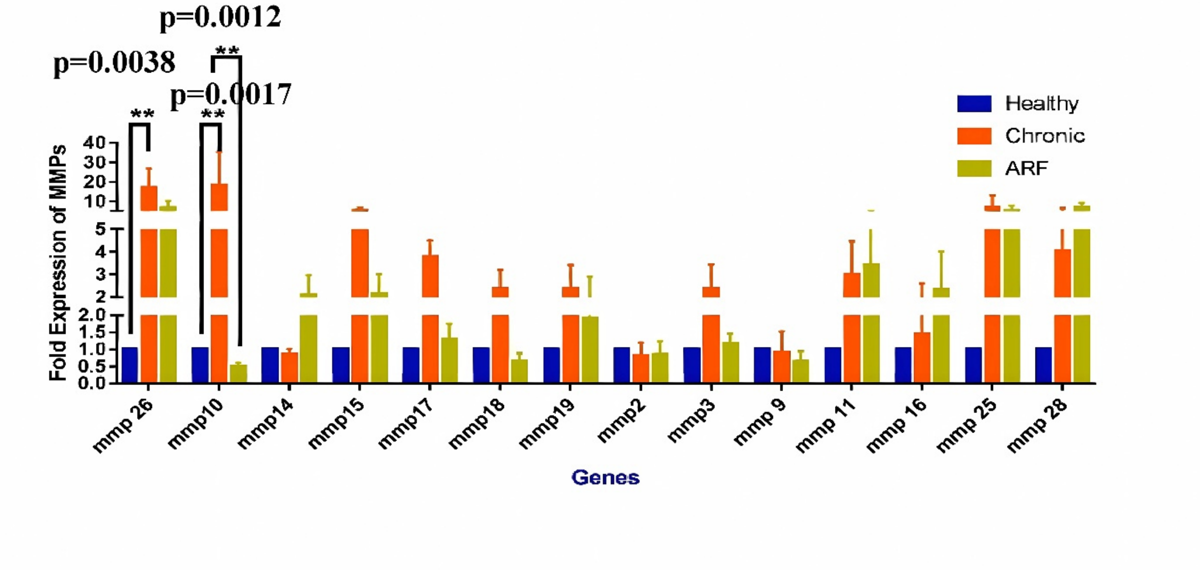


*Figure S2. Gene expression of Matrix metalloproteinases (MMPs) in the whole blood. Data are mean ± SEM and analysis was done using two-way ANOVA with Tukey's test for multiple comparasion. .*; p<0.05. n= ARF=3 Chronic=3, Healthy=3.The significantly upregulated expression of MMP-10 in total blood was selected.*

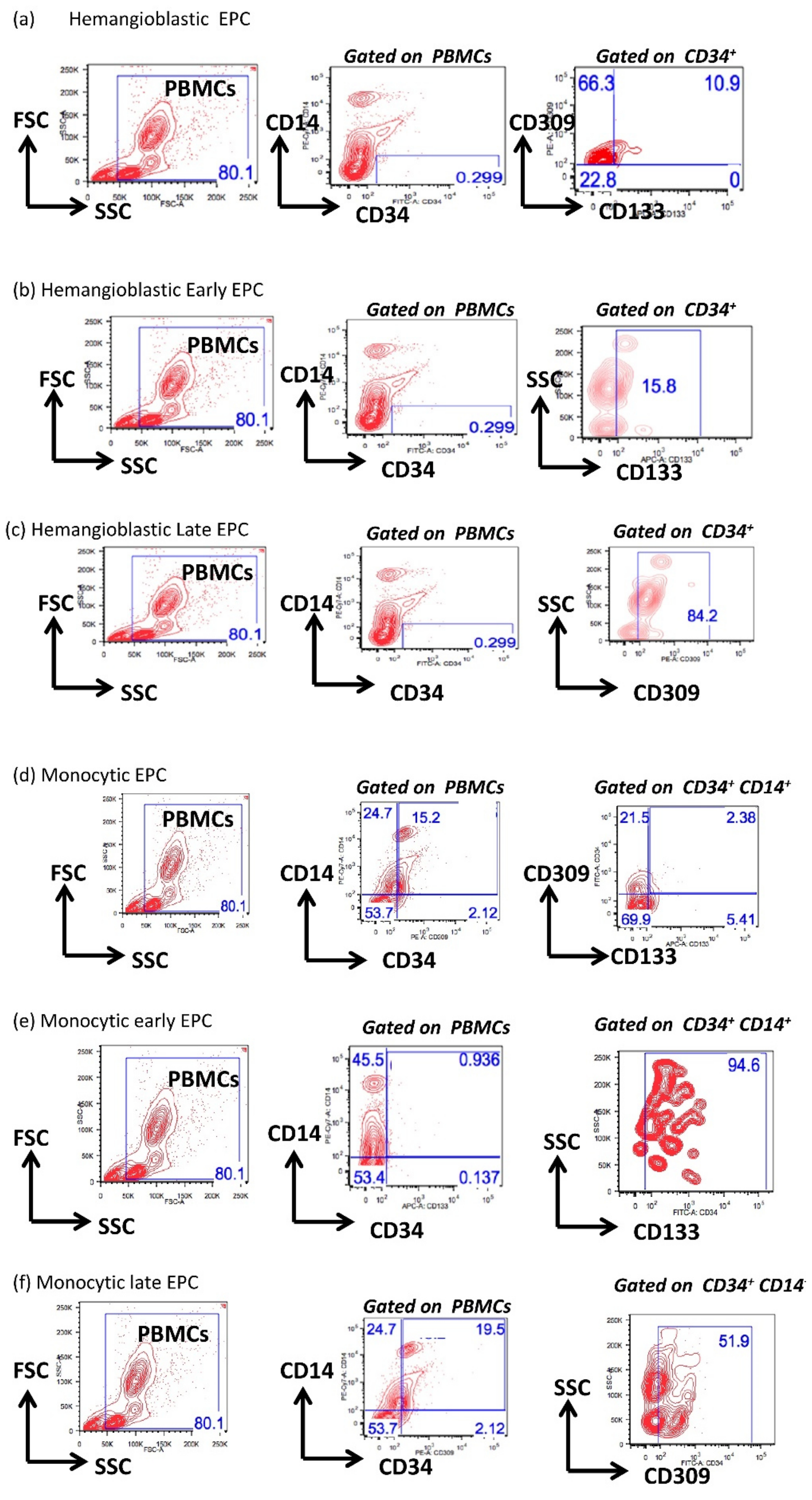


*Figure S3. Gating strategy for all the EPCs subsets and a representative graph. Panel (i) of all subsets from a to f represents the flow cytometer gating strategies used for analyzing the different subsets of EPCs. Panel (ii) is the respective graphs obtained after the enumeration of different types of EPCs. All the data are represented in median with interquartile range.* p<0.05. n=32 (ARF) n=20 (RHD), n= 15 (healthy controls).*

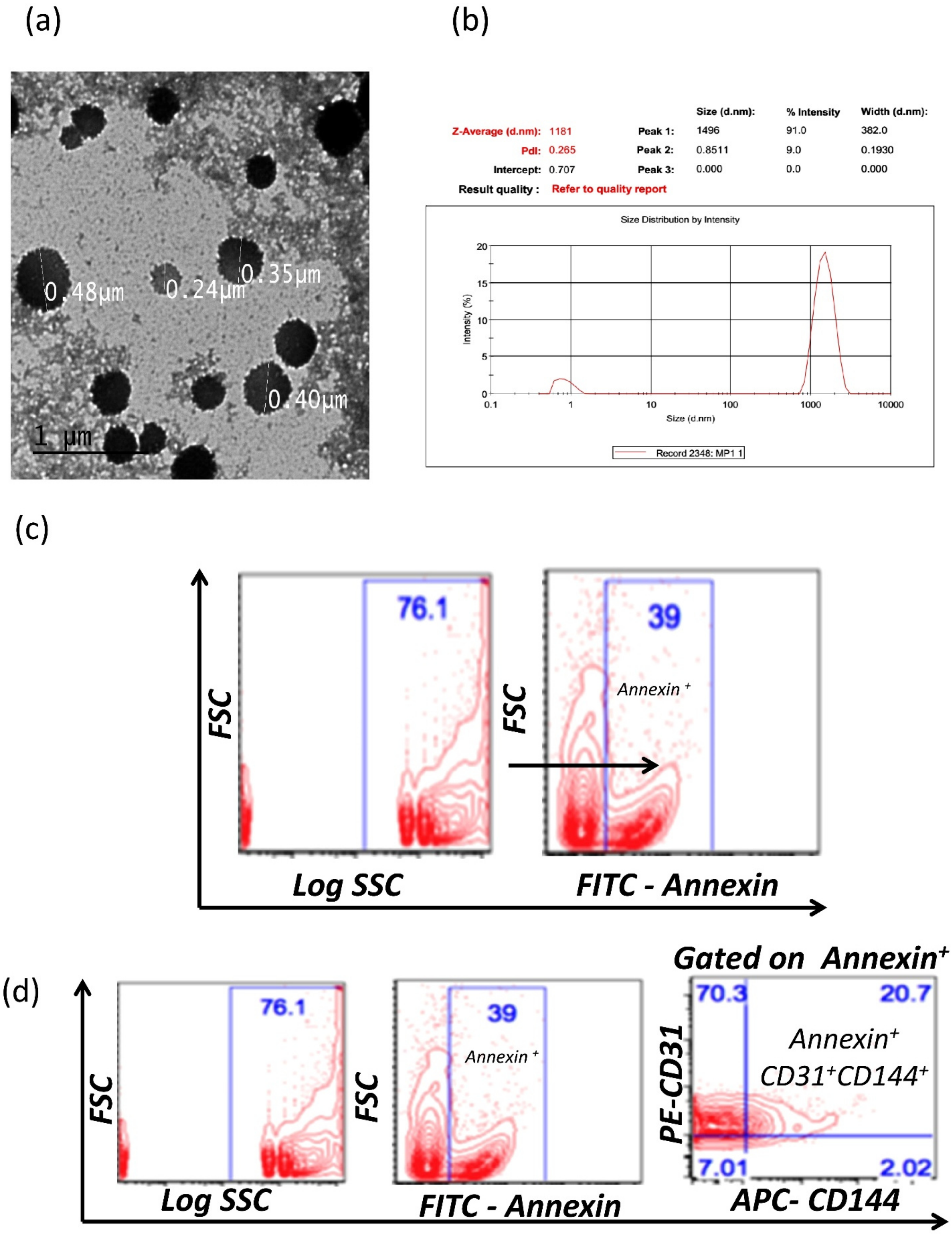


*Figure S4. Size, morphology and percentage of microvesicles. (a) an image of microvesicles by transmission electron microscopy (TEM) and (b) size distribution by dynamic light spectroscopy (DLS), (c) The isolated microvesicles were stained for Annexin$^{+}$, (d) The isolated microvesicles were also stained for endothelial derived microvesicles (Annexin$^{+}$ CD144$^{+}$ CD31$^{+}$).*

**Enumeration of monocyte microvesicles with their specific cell types.**

Therefore, monocyte (CD14$^+$) cells and monocyte progenitor cells (CD14$^+$CD34$^+$) cells were enumerated in the PBMCs of ARF, chronic and healthy subjects. These cells were then compared with their respective microvesicles, monocyte microvesicles (Annexin$^+$ CD14$^+$) and monocyte progenitor microvesicles (Annexin$^+$ CD14$^+$ CD34$^+$).

Circulating monocytes (CD14$^+$) and monocyte progenitor cells (CD14$^+$CD34$^+$) were enumerated in PBMCs of ARF, chronic and healthy subjects. PBMCs were stained with anti-human CD14-pecy7A and anti-human CD34-FITC.

Microvesicles were isolated from the plasma of ARF, chronic and healthy subjects. These MPs were stained using anti-human annexin-FITC, anti-human CD14-V421 and anti-human CD34-PEcy7A. Monocyte microvesicles (Annexin$^+$ CD14$^+$) and monocyte progenitor cell microvesicles (Annexin$^+$ CD14$^+$ 34$^+$) were enumerated.

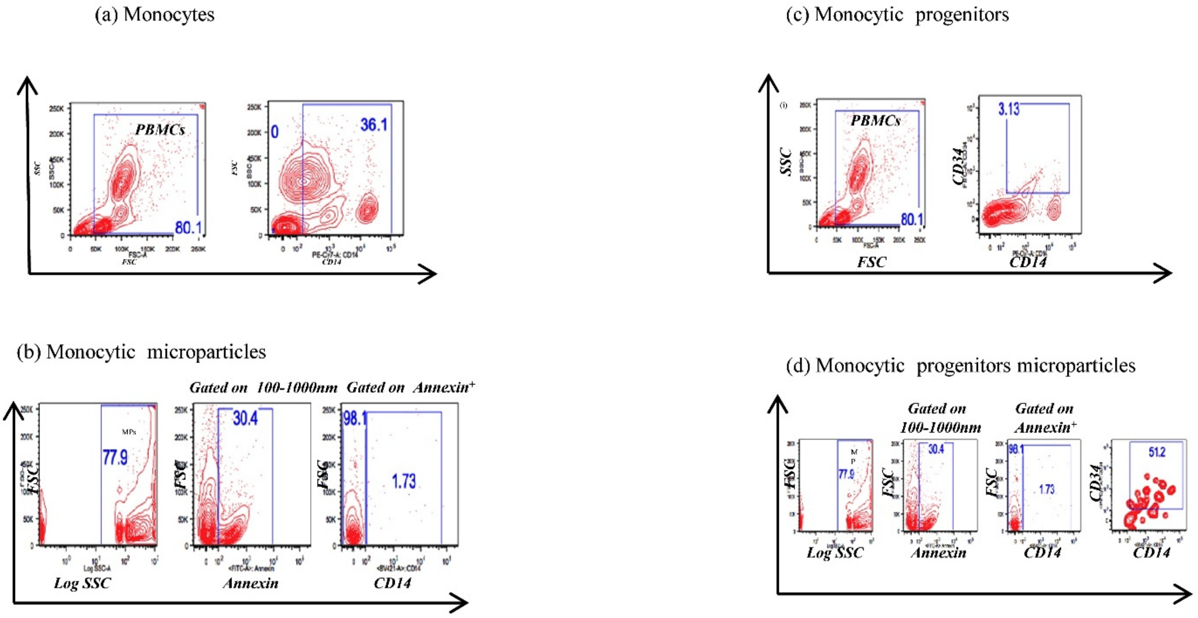


Figure S5. The comparison of all the monocyte and their subtypes,

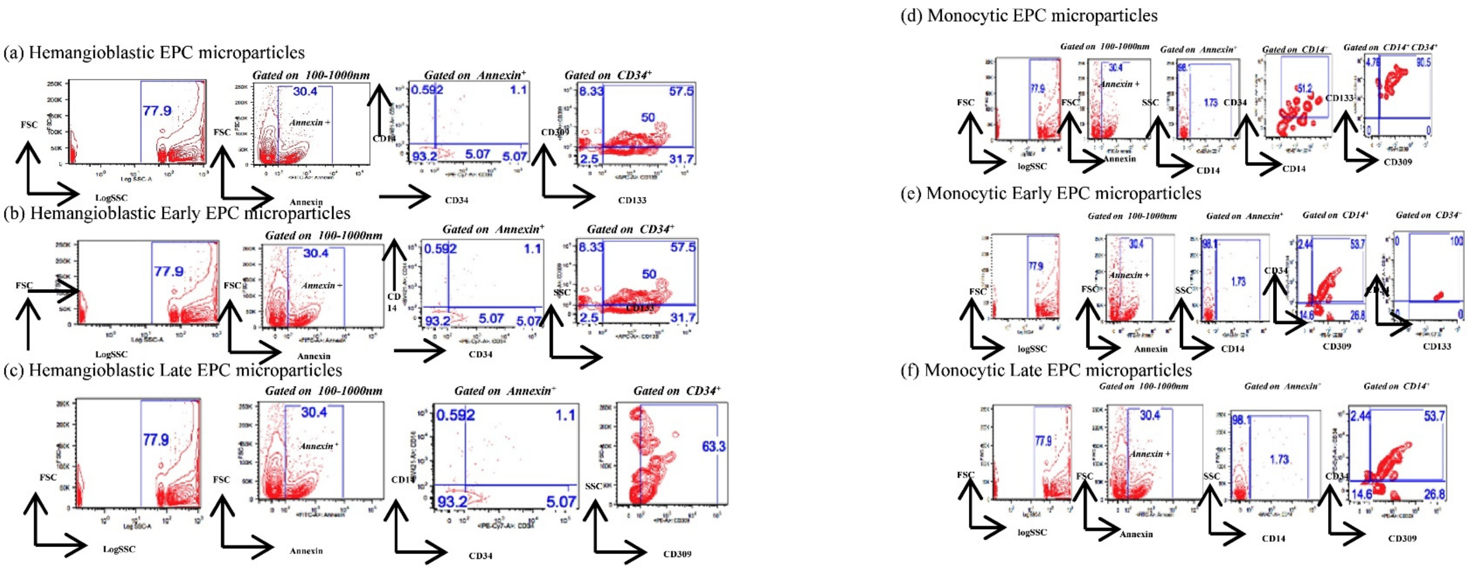


*Figure S6. Gating strategy for all the EPCs microparticles.*
